# Maternal Serum Hemoglobin Levels Relative to Anemia Interventions in the COVID-19 era: A systematic review and *meta*-analysis

**DOI:** 10.1101/2025.01.15.25320586

**Authors:** John K. Muthuka, Muthoni L. Agnes, Ngeno K. Leonard, Chembugei C. Lucy, Mbari F. Dianna, Rosemary Nabaweesi

## Abstract

**Aims:** The purpose of this review was to investigate the effect of different types of maternal anemia interventions on serum hemoglobin levels among pregnant cohort of reproductive age during COVID-19 pandemic using a pool of studies conducted with different research designs.

**Methods:** Relevant studies were identified from January 2020 to December 2022 by using MeSH terms in PubMed, Embase, Cochrane Library, Clinical trials, Scopus databases and gray literature. The studies were systematically reviewed, and quality assessments were evaluated by the guidelines of the Cochrane risk of bias. All the statistical analysis was performed on RevMan 5.4.1 software with a random and fixed effect models. Heterogeneity was evaluated with the Cochran Q statistic and Higgins test and publication bias assessed via funnel plots. The cumulative pooled effect of maternal anemia interventions (*n* = 15) on serum hemoglobin concentration was the main outcome, while effects of specific types of maternal anemia interventions; mode of administration; and the country a study was conducted were derived as secondary outcomes. In total, 6695 pregnant women of reproductive age were established from the entire pool of studies. Mean serum hemoglobin levels’ data were retrieved alongside their standard errors for ascertaining the effect of the said maternal anemia interventions on hemoglobin levels using f standard mean and mean differences during meta-analysis. The review component was part of registered PROSPERO: CRD-CRD42023410657:[

**Results:** Fifteen studies met the inclusion criteria, with the current study findings proposing that, maternal anemia interventions in overall improved serum hemoglobin levels [SMD (95% CI) = 0.93 [0.57, 1.30]; Heterogeneity: Tau-squared = 0.50; Chi-squared = 627.07, df = 14 (P < 0.00001); I-squared = 98%. The effect of specific intervention categories was; dietary iron supplement 1.16 [95% CI = −0.97, 3.28], education information 0.75 [95% CI = 0.12, 1.37], Intravenous ferric carboxy-maltose 1.33 [95% CI = 0.28, 2.38], intravenous iron sucrose 0.82 [95% CI = 0.56, 1.09] and herbal substances 0.42 [95% CI = 0.24, 0.61]. Dietary iron supplementation didn’t demonstrate an improvement on hemoglobin levels (Test for overall effect: Z = 1.07 (P = 0.29). Parenterally given and via education administered interventions had better utility on hemoglobin concentrations with maternal anemia interventions among developing countries showing insignificant effect on maternal serum hemoglobin levels.

**Conclusion:** Current review concluded that maternal anemia interventions had a utility on serum hemoglobin, more feasible, those given as injections. Notably, dietary iron interventions were compromised especially in developing nations during COVID-19 pandemic. Multifaceted further clinical studies are crucial to substantiate current findings.

## Introduction

Maternal anemia is a significant global health concern affecting a large number of women, particularly those in low- and lower-middle-income countries. Around 37% of pregnant women aged 15-49 years being affected. This translates to millions of women worldwide[1]. Anemia during pregnancy is associated with poor maternal and birth outcomes, such as premature birth, low birth weight, and maternal mortality[2]. In many regions, especially low- and middle-income countries, access to prenatal care is limited. This means that many pregnant women do not receive the necessary screenings and interventions to prevent or treat anemia[3], [4]. Limited resources and funding can hinder the implementation of effective anemia prevention and treatment programs. This includes shortages of iron supplements, nutritional education, and healthcare personnel[5].

The COVID-19 pandemic has had a profound impact on global health services and interventions across the board and has significantly impacted maternal anemia interventions[6], [7]. Lockdowns, curfews, and movement restrictions made it difficult for patients to access healthcare facilities. Many people avoided hospitals and clinics for fear of contracting the virus[8]. Many pregnant women missed routine prenatal check-ups due to lockdowns and fear of contracting the virus[9]. The scoping review provides a comprehensive overview of both the direct and indirect effects of the pandemic on maternal and perinatal health highlights pregnant women were found to be at a heightened risk of more severe symptoms compared to non-pregnant individuals[9].

Interventions to address maternal anemia at a global level are multifaceted and involve various strategies to improve the health and well-being of women of reproductive age. Routine supplementation with iron and folic acid (IFA) is a cornerstone intervention[10]. Pregnant women are often advised to take 60 mg of iron and 400 mcg of folic acid daily. Fortifying commonly consumed foods with essential vitamins and minerals can help address nutritional deficiencies and encouraging a diet rich in iron-rich foods such as lean meats, fish, beans, and leafy green vegetables[11], [12]. Intravenous ferric carboxy-maltose (FCM) is a promising intervention for treating iron deficiency anemia in pregnant women showing that it significantly increases hemoglobin levels in iron deficiency anemia[13]. This improvement is observed within a few weeks of administration[14].

The World Health Organization (WHO) has conducted extensive research and published several studies on maternal anemia interventions, particularly focusing on the differences between developing and developed countries[15]. There is a framework outlining strategies to accelerate the reduction of anemia in women of reproductive age. It emphasizes the importance of addressing both direct causes (like micronutrient deficiencies) and underlying risk factors (such as poverty and gender inequities)[16]. Notably, it has mapped evidence on interventions targeting anemia’s direct causes and underlying risk factors. This includes nutritional interventions, healthcare improvements, and policy advocacy[17]

In developed countries, anemia management often involves routine prenatal care, widespread use of iron and folic acid supplements, and advanced healthcare infrastructure. In contrast, developing countries face challenges such as limited access to healthcare, higher prevalence of infectious diseases (like malaria), and socioeconomic barriers. Interventions in these regions focus on improving healthcare access, nutritional education, and addressing broader social determinants of health1[15], [18].

This systematic review and meta-analysis of RCTs, quasi experiments and observational studies is carried out to endeavor summarizing the evidence on the effects of different types of maternal anemia interventions on serum hemoglobin levels using majorly SMD with random effects modelling. Further, it aims at evaluating the heterogeneity among the said pooled studies’ results in pregnant subjects of reproductive in the advent of COVID-19.

## Materials and methods

### Design

All guidelines listed in the Preferred Reporting Items for Systematic Reviews and Meta-Analyses (PRISMA) statement were adhered to in conducting this meta-analysis [19]. Notably, the PRISMA is primarily used for preparing systematic reviews of such research interventions [31, 32]. Data were pooled from random clinical trial (RCTs), observational, including cohort, case-control, cross-sectional, and similar viable case studies. The study was PROSPERO registered (CRD-CRD42023410657).

### Search Strategy

The search terms were carried out in the PubMed, Embase, Cochrane Library, and Scopus databases by two independent investigators, and relevant publications cited in English from December 2019 to August 2022 using search the following terms: “maternal anemia” OR “anemic condition” OR “hemoglobin levels)” OR “pregnancy anemia” OR “anemia in pregnant women” OR “gestation anemia” OR “hemoglobin concentration” OR “hemoglobin changes” OR “hemoglobin status” AND “treatment” OR “intervention” OR “pregnancy anemia management” AND “effect” OR “effectiveness” AND “impact” OR “outcome”. The titles and abstracts of the screened articles were checked, and duplicate citations were then removed. After excluding non-relevant articles, full texts of the selected articles were retrieved considering only articles in English.

### Study selection

#### Inclusion criteria

A structured approach was taken to set up the research question regarding this current review, using the following five components commonly known as the Participants, Interventions, Comparisons, Outcomes, and Study Design Approach (PICOS) [33]:

i. studies that examined women within reproductive age and put on any anemia prevention program or intervention, either anemic or non-anemic according to World Health Organization (WHO) criteria in the advent of COVID-19 pandemic.
ii. Studies reporting the effects of any feasible form of maternal anemia interventions on hemoglobin levels, status, change or concentration as primary or secondary outcomes were considered. No restrictions were placed on the gender, age, race, and geographical distribution of the individuals enrolled in the study.
iii. Maternal anemia interventions such as; nutrition education, dietary iron supplement, intravenous ferric-carboxyl maltose administration, intravenous iron sucrose administration, any non-convectional maternal anemia management remedy and any other possible intervention.
iv. Study design: both RCTs and Observational studies including; cohort, cross-sectional, case-control and also, quasi experiments

#### Exclusion criteria

i. Editorials, case reports, letters to the editor, review articles, and animal studies.
ii. RCTs or observational studies not evaluating hemoglobin levels or concentrations or changes or status as their primary or secondary outcomes; or studies with no controls or comparator interventional arm.
iii. RCTs or observational studies that did not report mean (SD) and mean values hemoglobin, levels in intervention or control/comparator groups.
iv. Non-English-language studies lacking a full text (unavailable or not yet published), without a DOI number and with small sample sizes (<50 patients) because of low statistical power.

### Data Extraction

Both adjusted and non-adjusted data among pregnant women on experimental interventions versus pregnant women on comparators intervention (control) were extracted to identify the most relevant confounding factors to be used in the analysis by subsequent pooling. Two researchers completed the data extraction independently (JK and AM) where qualitative and quantitative information were extracted from each study. Any disagreement between the researchers with respect to the inclusion criteria of a study was resolved by the insight of a third researcher (LN). Only studies agreed upon by all reviewers were included in the final analysis. The following data were obtained from all studies: title, first author’s name, data collection year, region, sample size, study design, study setting (single or multicenter), maternal anemia intervention type, and the hemoglobin mean values alongside their associated standard deviations (SD) as either the primary or secondary outcome and conclusions. *P*-value of < 0.05 was considered statistically significant for all of the included studies.

### Quality and risk of bias assessment

The risk of bias was assessed using the Cochrane risk of bias tool, and the RoB 2 tool (7.0) [20]. The included RCTs were assessed in the following domains: risk of bias arising from the randomization process, risk of bias due to deviations from the intended interventions, risk of bias due to missing outcome data, risk of bias in measurement of the outcome, and risk of bias in selection of the reported result, followed by the assessment of the overall risk of bias. On the other end, the National Institutes of Health tool for observational studies [21] was used for methodological quality assessment. Two to three reviewers independently assessed the quality of the studies, and the scores were added to the data extraction form before inclusion in the analysis to reduce the risk of bias. To evaluate the risk of bias, the reviewers rated each of the 14 items into qualitative variables: yes, no, or not applicable. An overall score was by adding the scores of all items with yes=1 and no or not applicable=0 was performed. A score classification of poor (score 0-5), fair (score 6-9), or good (score 10-14) was given for each study. Data were checked by reviewers who did not perform the data extraction or each reviewer was assigned an article that they had not extracted data from in previous steps.

### Statistical analysis

In the current review, all the statistical analysis was performed on RevMan 5.4.1 software with a random and fixed effect model. Random effect model was used for random variances, when the number of studies entered was limited and there was a difference between the number and characteristics of the individuals (DerSimonian R, Laird N. Meta-analysis in clinical trials. Control Clin Trials). Small and large sample sizes had the same effect in the final conclusion when using this model. The fixed effect model was used for studies with fixed parameters or non-random quantities. Continuous data from all the studies was extracted and accumulated, then the variables were analyzed in order to obtain overall hemoglobin level mean difference (MD) with 95% confidence intervals (CIs) by the inverse variance approach. The effect of maternal anemia interventions on mean hemoglobin levels were compared between the intervention and the comparator/control arms. Heterogeneity was evaluated with the Cochran Q statistic and Higgins test. The Higgins test uses a fixed-effects model when the heterogeneity is <50% and a random-effects model when the heterogeneity is >50% (Cochrane Handbook for Systematic Reviews of Interventions | Cochrane Training). When heterogeneity was detected, a sensitivity adjustment and sub-group analysis was made to determine its source. It was considered statistically significant if *P* value was less than 0.05. Publication bias was evaluated using the Cochrane Risk of Bias tool.

### Subgroup analysis

Predetermined subgroup analyses were performed according to Higgins [22], to evaluate the potential effects of different maternal anemia interventions on hemoglobin levels among pregnant women in the advent of COVID-19. This method, studies were explored according to the potential heterogeneity of inducer factors, thus separate statistical analyses were performed in each study subgroups. Thereafter, studies were categorized according to the different maternal anemia interventions, and then separate meta-analysis was conducted. A significant reduction in the extent of heterogeneity in each subgroup, confirmed the heterogeneity based on the different maternal anemia interventions. Studies were further categorized based on study design, year(s) data was collected and the study setting.

## Results

PRISMA flow diagram illustrates the selection of included studies and screening process in this current review. In total, 460 articles were found in the initial search, and 445 of these articles were excluded after reading the titles and abstracts where supplementation of interest was not evaluated. Additionally, duplicates were removed. Finally, 15 studies [23], [24], [25], [26], [27], [28], [29], [30], [31], [32], [33], [34], [35], [36], [37] met the inclusion criteria and were suitable for quantitative synthesis (Fig. 1)

**Figure 1.**
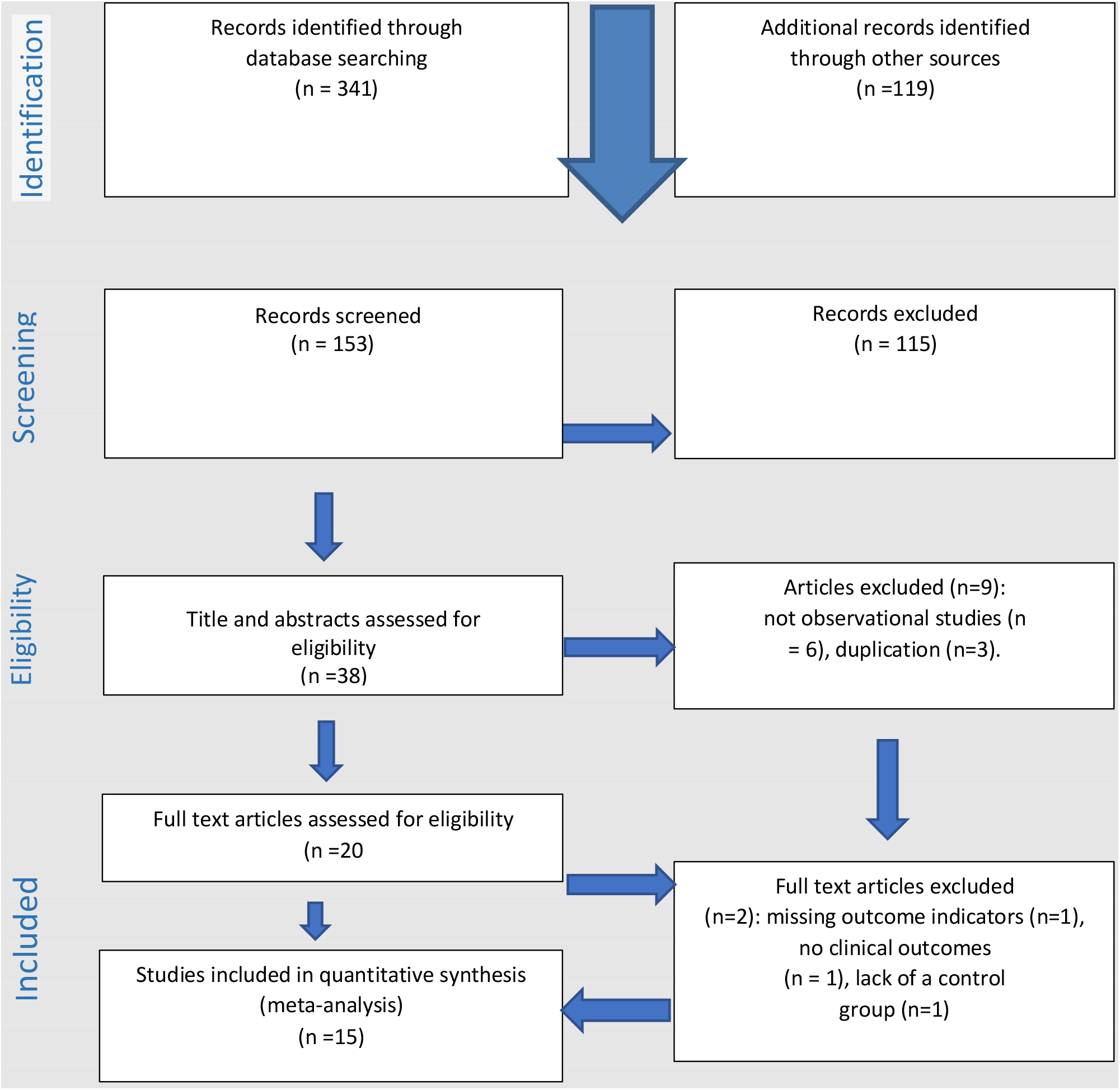
PRISMA flow-diagram of the study selection process

### Study characteristics

Most of the studies with a reservation for four [24], [25], [34], [37] were experimental in design with two [27], [32] being quasi experiments. The main characteristics of the studies are illustrated in Table 1. Studies were published online between 2021 and 2023. The range of sample size was from 110 to 1644 participants in terms of the total sum between the intervention and the control arm. Close to 50% of the studies included here showed education package and or some form of information rendered to pregnant women as the most common form of intervention then with majority having being conducted from Asian and African countries.

### Maternal anemia Intervention characteristics

Different types of interventions were used in these studies, in seven studies, the intervention was in form of an education and or information package[23], [27], [28], [29], [30], [31], [32], in four, the intervention was ferric carboxy-maltose [24], [33], [34], [35], one, Dietary iron intake [25], one, intravenous iron sucrose group [36], one, IFA + micronutrient-fortified BEP supplement [26] and one, a fresh moriga consumption [37]. Generally, the interventions were given between the year 2020 to the year 2022 (Table 1).

### Outcome measures

All the 15 studies reported hemoglobin levels changes post maternal anemia intervention alongside their standard deviations as the primary outcomes without any further consideration of other biomarkers such as iron markers including; levels of ferritin, serum iron, and transferrin secondary outcomes. The lowest and highest hemoglobin level post-intervention in intervention arm was 10.12(g/dL) [32] and 12.77(g/dL)[29] respectively, while in control or comparator arm, the lowest and highest was 9.43(g/dL) [25] and 13.09 (g/dL)[29] respectively (Table 1).

**Table 1.** Studies on maternal anemia interventions effect on hemoglobin levels in the advent of covid-19.

|  |  |  |  |  |  |  |
| --- | --- | --- | --- | --- | --- | --- |
| 1) | [25] | Africa | <sup>g</sup>CS, <sup>b</sup>MC | Dietary iron intakes | 2020 |  |
| 2) | [27] | Asia | <sup>c</sup>RCT, <sup>h</sup>SC | Nutrition Education | 2020 | – |
| 3) | [29] | Asia | <sup>i</sup>Q-E, <sup>h</sup>SC | The educational intervention | 2021 |  |
| 4) | [36] | Asia | <sup>c</sup>RCT, <sup>h</sup>SC | Intravenous iron sucrose group | 2020–2021 |  |
| 5) | [28] | Asia | <sup>c</sup>RCT, | Individual education was delivered on the HIPPI | 2021 |  |
| 6) | [26] | Africa | <sup>f</sup>RCT, <sup>b</sup>MC | IFA + micronutrient-fortified BEP supplement | 2021 |  |
| 7) | [34] | Asia | <sup>c</sup>RCC, <sup>h</sup>SC | Intravenous ferric carboxy-maltose treatment | 2020 |  |
| 8) | [30] | Africa | <sup>c</sup>RCT, <sup>b</sup>MC | Nutrition Information | 2020-2021 |  |
| 9) | [35] | Africa | <sup>c</sup>RCT, <sup>b</sup>MC | Ferric carboxy-maltose | 2018-2021 |  |
| 10) | [32] | Asia | <sup>i</sup>QE, <sup>h</sup>SC | Individual nutrition education | 2021 |  |
| 11) | [24] | Europe | <sup>k</sup>C C, <sup>h</sup>SC | Intravenous ferric carboxy-maltose | 2021A |  |
| 12) | [31] | Asia | <sup>c</sup>RCT, <sup>b</sup>MC | MyPinkMom intervention Program-Education | 2020-2021 |  |
| 13) | [33] | Asia | <sup>c</sup>RCT, <sup>h</sup>SC | Intravenous ferric carboxymaltose | 2020 |  |
| 14) | [23] | Africa | <sup>c</sup>RCT, <sup>b</sup>MC | Intensive nutrition education | 2021-2022 |  |
| 15) | [37] | Africa | <sup>g</sup>CS, <sup>b</sup>MC | Moriga Leaf, Herbal | 2022 |  |

*aRCC: retrospective case-control. bMC: multicenter. cRCT: random clinical trial. dR: retrospective. eO: observational. fP: prospective. gCS: cross-sectional. hSC: single-center. iQE: quasi experiment.*

### Quality and risk of bias assessment

We assessed the quality of the included studies based on a modified version of the Newcastle-Ottawa Scale (NOS), which consists of 8 items with 3 subscales, and the total maximum score of these 3 subsets is 9. We considered a study that scored ≥7 to be a high-quality study since a standard criterion for what constitutes a high-quality study has not yet been universally established. The 15 studies assessed generated a mean value of 6.8, indicating that the overall quality was moderate (NOS score range 5-8) (Table 2).

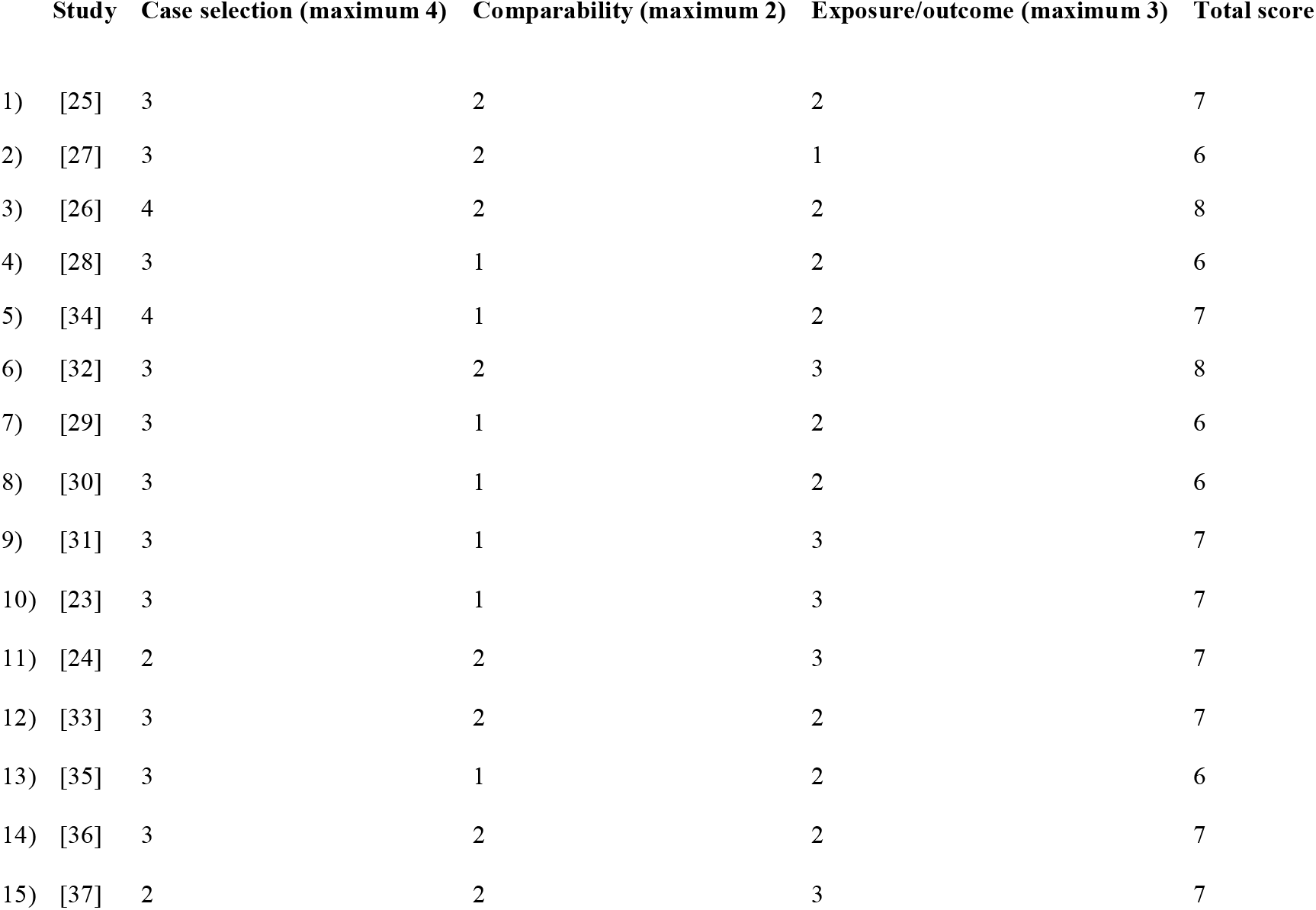

| Study | Case selection (maximum 4) | Comparability (maximum 2) | Exposure/outcome (maximum 3) | Total score |
| --- | --- | --- | --- | --- |
| 1) [25] | 3 | 2 | 2 | 7 |
| 2) [27] | 3 | 2 | 1 | 6 |
| 3) [26] | 4 | 2 | 2 | 8 |
| 4) [28] | 3 | 1 | 2 | 6 |
| 5) [34] | 4 | 1 | 2 | 7 |
| 6) [32] | 3 | 2 | 3 | 8 |
| 7) [29] | 3 | 1 | 2 | 6 |
| 8) [30] | 3 | 1 | 2 | 6 |
| 9) [31] | 3 | 1 | 3 | 7 |
| 10) [23] | 3 | 1 | 3 | 7 |
| 11) [24] | 2 | 2 | 3 | 7 |
| 12) [33] | 3 | 2 | 2 | 7 |
| 13) [35] | 3 | 1 | 2 | 6 |
| 14) [36] | 3 | 2 | 2 | 7 |
| 15) [37] | 2 | 2 | 3 | 7 |

### Meta-analyses

#### Outcomes

The data from fifteen studies [23], [24], [25], [26], [27], [28], [29], [30], [31], [32], [33], [34], [35], [36], [37] examined the effect of different maternal anemia interventions on hemoglobin levels as the key biomarker used as an indicator of maternal anemia in the era of COVID-19 pandemic between intervention and comparative arms.

#### Cumulative effect of maternal anemia interventions on hemoglobin levels

Ten random clinical trials [23], [26], [27], [28], [30], [31], [33], [35], [36] (n = 5,295), two quasi experiments [29], [32] (n= 2435) and four observational studies [24], [25], [34], [37] (n = 1151) reported mean hemoglobin concentrations alongside their specific standard errors. On performing pooled analysis using random effects model, the maternal anemia interventions in overall improved hemoglobin levels with a significant heterogeneity existing in the data [SMD (95% CI) = 0.93 [0.57, 1.30]; Heterogeneity: Tau^2^ = 0.50; Chi^2^ = 627.07, df = 14 (P < 0.00001); I^2^ = 98%. We categorized our data in to five groups based on the type of intervention via sub group analysis which demonstrated a pooled effect of dietary iron supplement 1.16 [95% CI = −0.97, 3.28]− −0.97]; I^2^ = 99%, education information 0.75 [95% CI = 0.12, 1.37]; I^2^ = 97%, I.V ferric carboxy-maltose 1.33 [95% CI = 0.28, 2.38]; I^2^ = 99%, intravenous iron sucrose 0.82 [95% CI = 0.56, 1.09]; I^2^ = not applicable and herbal/non-convectional substances 0.42 [95% CI = 0.24, 0.61]; I^2^ = not applicable and, was notable that, dietary iron supplementation among pregnant women during COVID-19 period didn’t demonstrate an improvement on hemoglobin levels (Test for overall effect: Z = 1.07 (P = 0.29) (Fig. 2).

**Figure 2.**
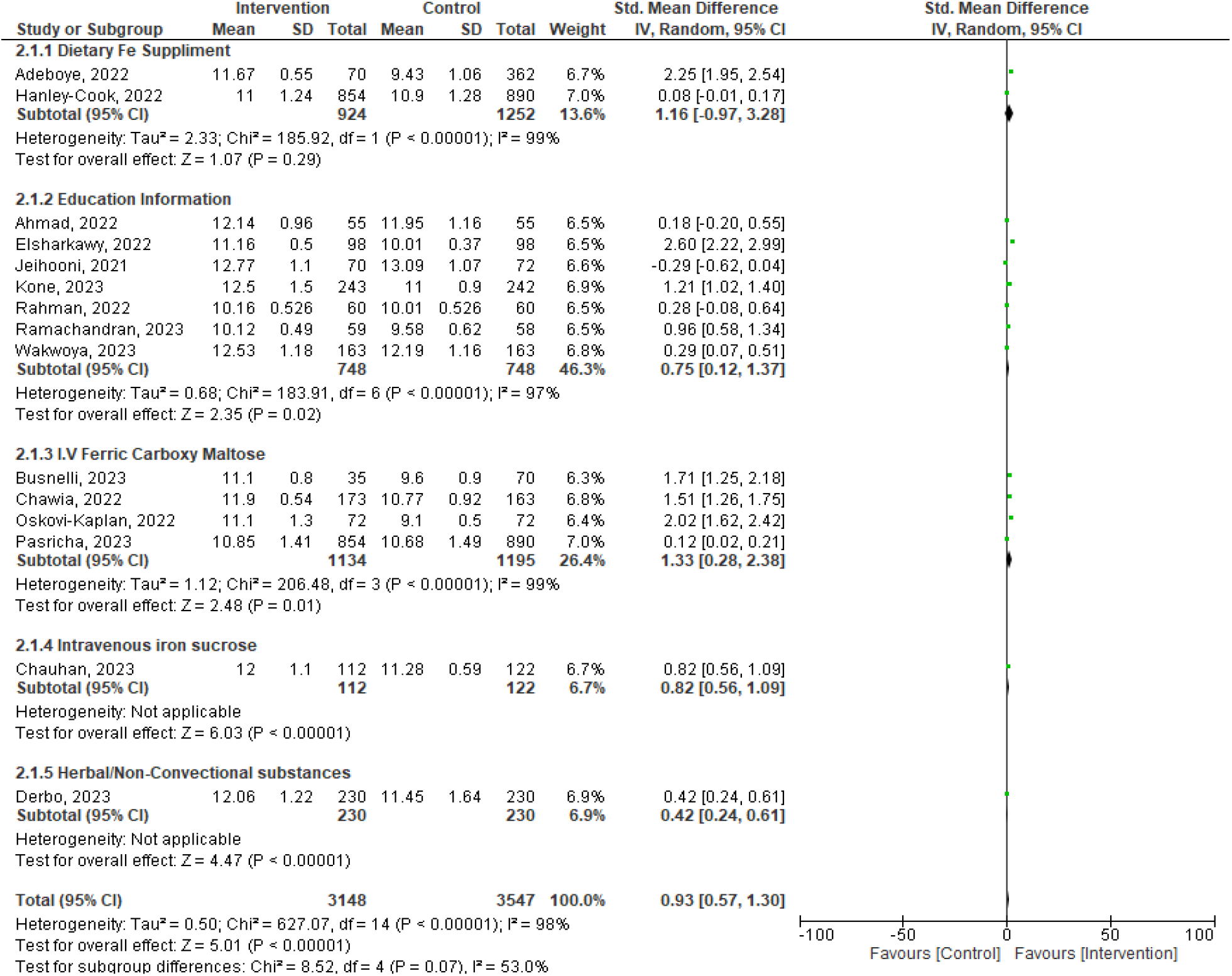
Forest plot showing results of a meta-analysis on the effects of maternal anemia interventions on hemoglobin levels. Data were reported as SMDs to compare the effect sizes across different studies to control for variability within the data, with 95% CIs.

#### A comparison between orally, parenterally and via education package administered maternal anemia interventions’ effect on serum hemoglobin levels

We clustered the studies based on the nature and or route at which the maternal anemia intervention was administered. The subgroup analysis showed that, parenterally given (injectable) interventions (n = 2563) majorly improved hemoglobin concentrations [MD (95% CI) = 1.09[0.48, 1.70]; p = 0.0004); Heterogeneity: Tau^2^ = 0.47; Chi^2^ = 167.61, df = 4 (P < 0.00001); I^2^ = 98% with orally administered interventions (n = 2,636) having no significant improvement on hemoglobin levels [MD (95% CI) = 0.98[-0.46, 2.43]; p = 0.0004); Heterogeneity: Tau^2^ = 1.61; Chi^2^ = 415.65, df = 2 (P < 0.00001); I^2^ = 100%. It was evident that, interventions given via education package or information (n=1496) had also, a significant improvement on hemoglobin levels 0.52 [95% CI = 0.07, 0.97]; I^2^ = 97%. A significant heterogeneity existed in the data. (Fig. 3).

**Figure 3.**
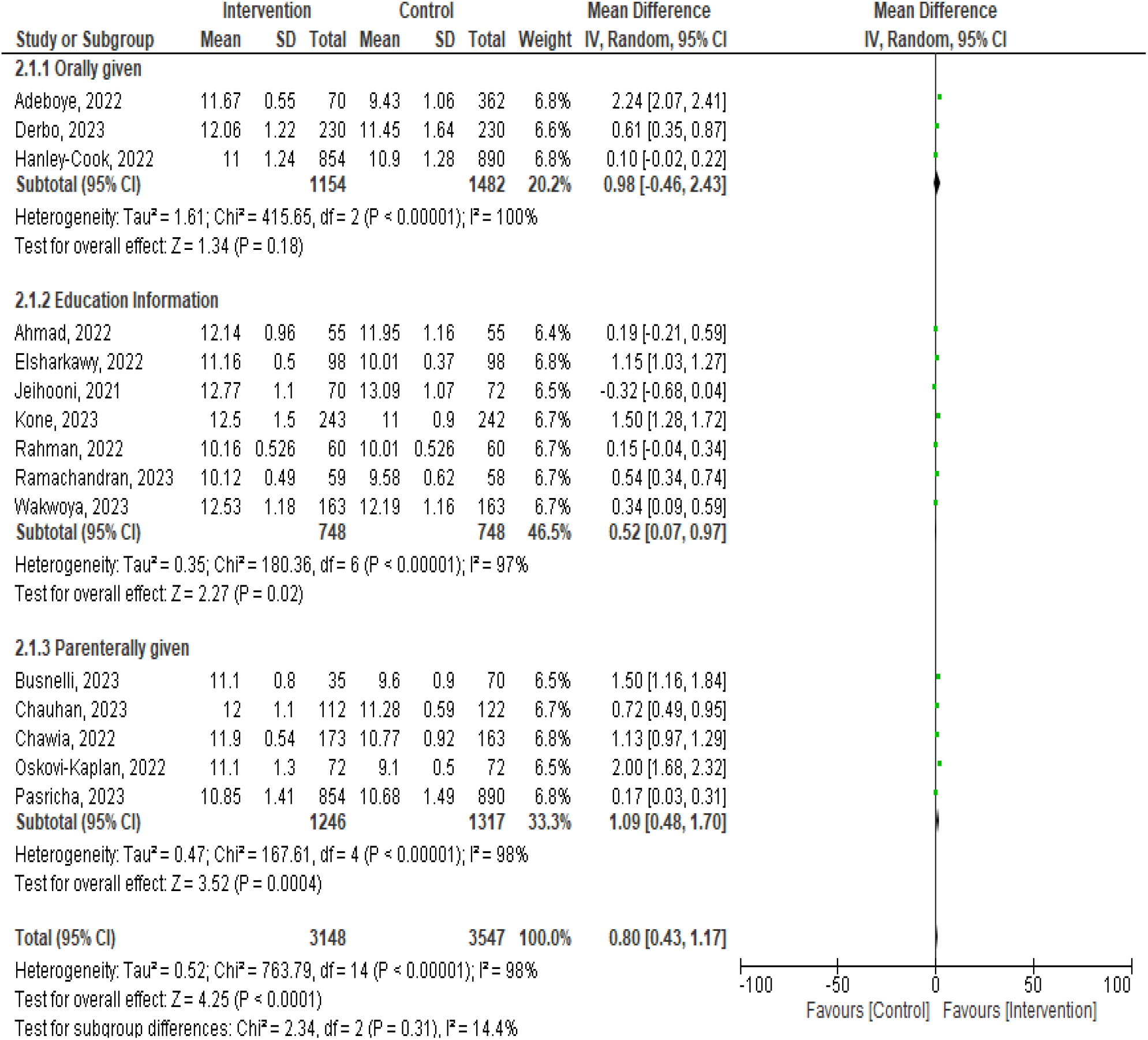
Forest plot showing results of a sub-group meta-analysis on the effects maternal anemia interventions on hemoglobin levels based on nature and or route of administration. Data were reported as MDs with 95% CIs.

With an assumption that the studies are sufficiently similar in terms of the interventions given subject to the mode and or route of administration (as being same), and there is no significant heterogeneity among them, a fixed-effect model was applied to robustly investigate the existing effect. The interventions orally given 0.76 [95% CI = 0.67, 0.84]; I^2^ = 97%, parenterally given 0.76 [95% CI = 0.67, 0.85]; I^2^ = 98% and via education package 0.76 [95% CI = 0.68, 0.84]; I^2^ = 97% basically showed a similar significant effect on hemoglobin levels with no heterogeneity in overalls relative the sub-groups [Test for subgroup differences: Chi^2^ = 0.16, df = 2 (P = 0.92), I^2^ = 0%] as demonstrated by the funnel plot (Figures 4 and 5).

**Figure 4.**
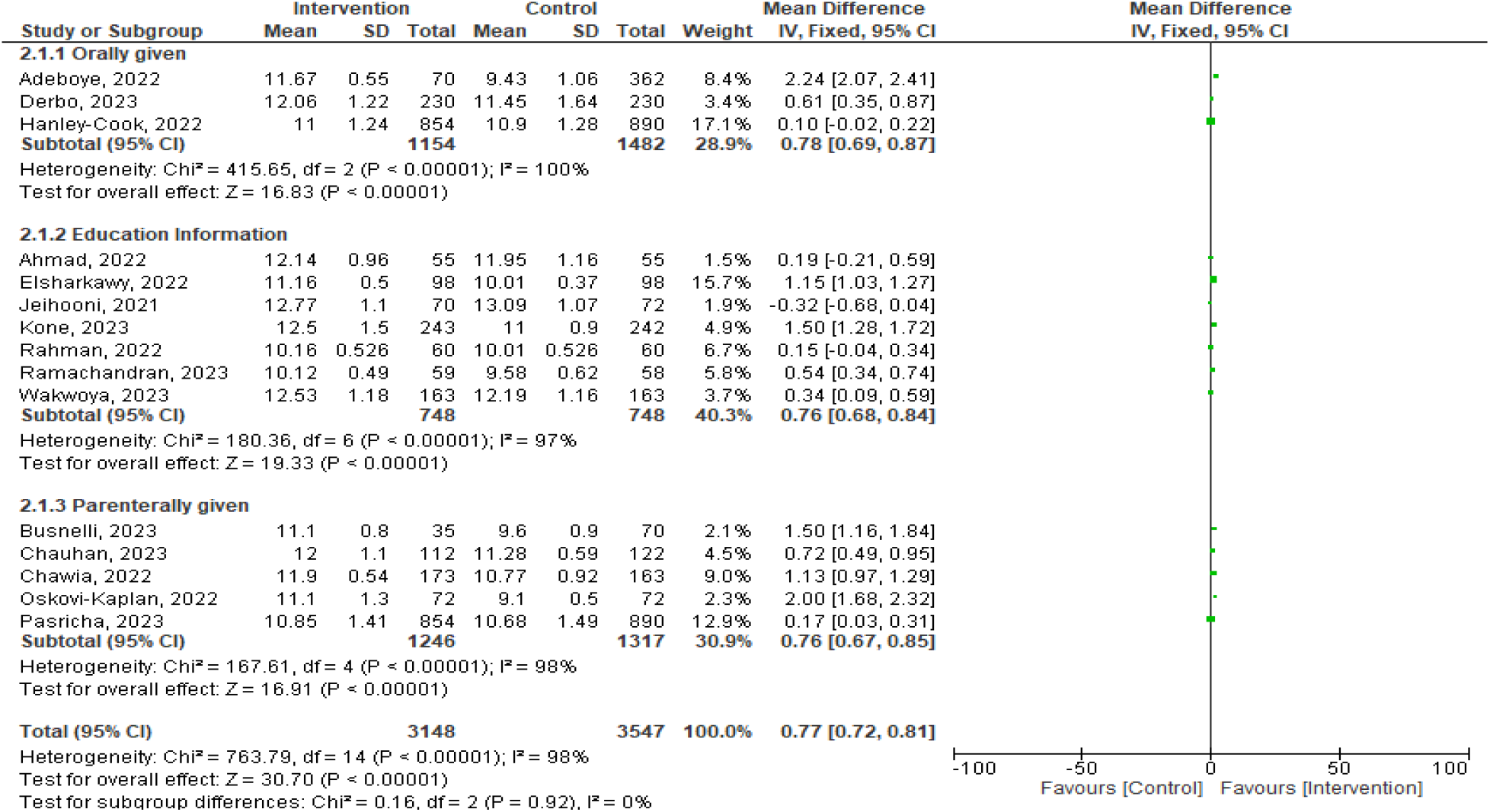
Forest plot showing results of a sub-group meta-analysis on the effects of maternal anemia interventions on hemoglobin levels based on the mode and or route of administration. Data were reported as MDs with 95% CIs.

**Figure 5.**
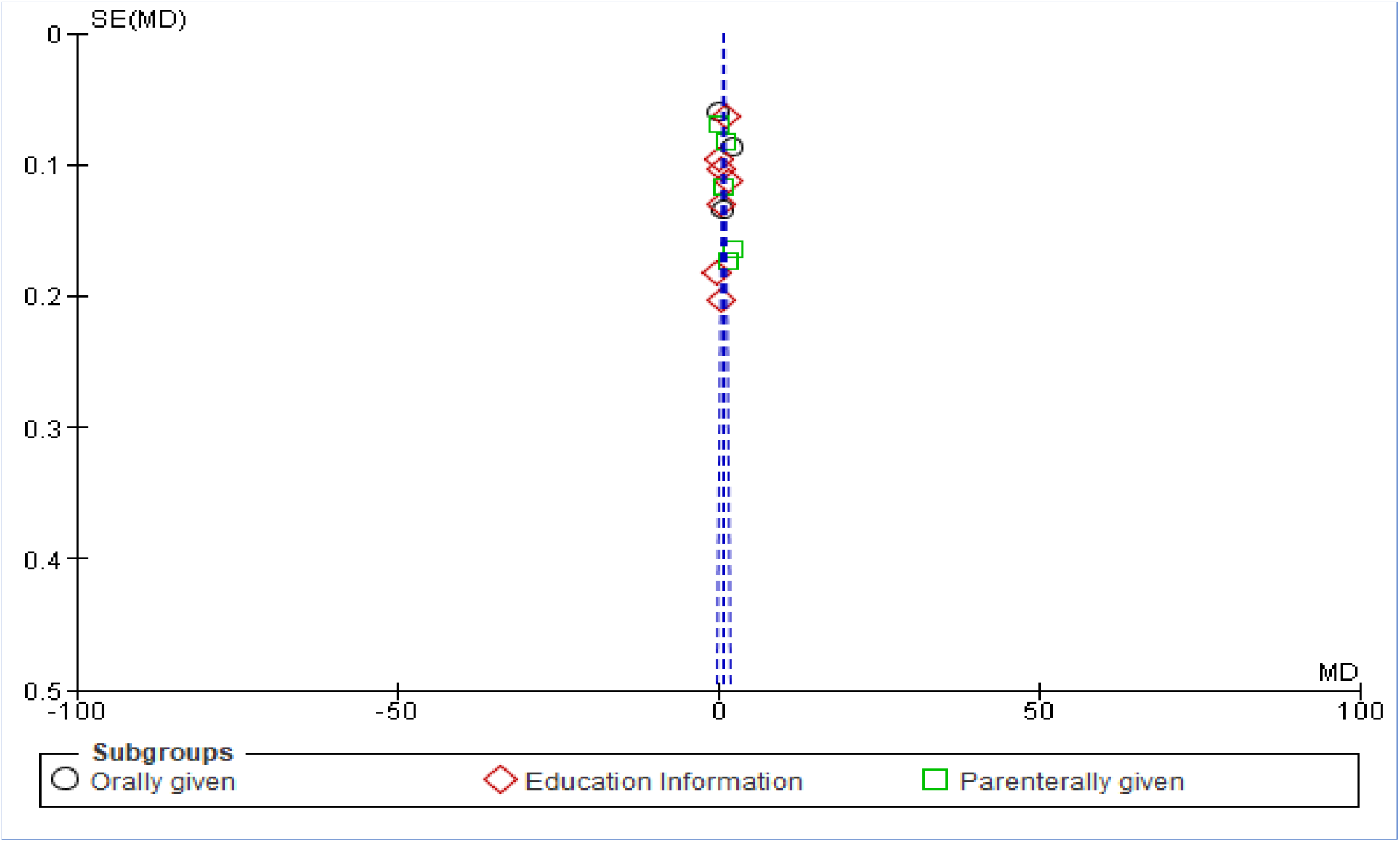
FA funnel plot showing results of a sub-group meta-analysis on the effects of maternal anemia interventions on hemoglobin levels based on the mode and or route of administration. Data were reported as MDs with 95% CIs.

#### Sensitivity analysis and publication bias

Sensitivity analysis using either; SMD or MD with either random effects or fixed effect models conducted via leave-one-out method did not cause a substantial change in the effect of specific maternal anemia interventions on hemoglobin levels. For instance, one study [25] removed, the effect of the intervention, the dietary iron intake did not significantly change the overall outcome of hemoglobin levels:[SMD (95% CI) = 0.84 [0.50, 1.18]; *p* < 0.0001); I^2^ = 97%] (n= 6,263) and another one [23] :[SMD (95% CI) = 0.98 [0.59, 1.37]; *p* = < 0.0001; I^2^ = 98%] (n=6,369), retaining a similar significance and effect as before the sensitivity analysis [SMD (95% CI) = 0.93 [0.57, 1.30]; Heterogeneity: Tau^2^ = 0.50; Chi^2^ = 627.07, df = 14 (P < 0.00001); I^2^ = 98%. Despite this, there was asymmetry observed (where studies tend to lie on the upper part of the overall effect line) showing publication bias and true heterogeneity (different populations or interventions leading to varying effects) seen in funnel plot Subject to this, current results showed that effect of specific maternal anemia interventions on hemoglobin concentrations had an evidence of publication bias. (Fig 6).

**Figure 6.**
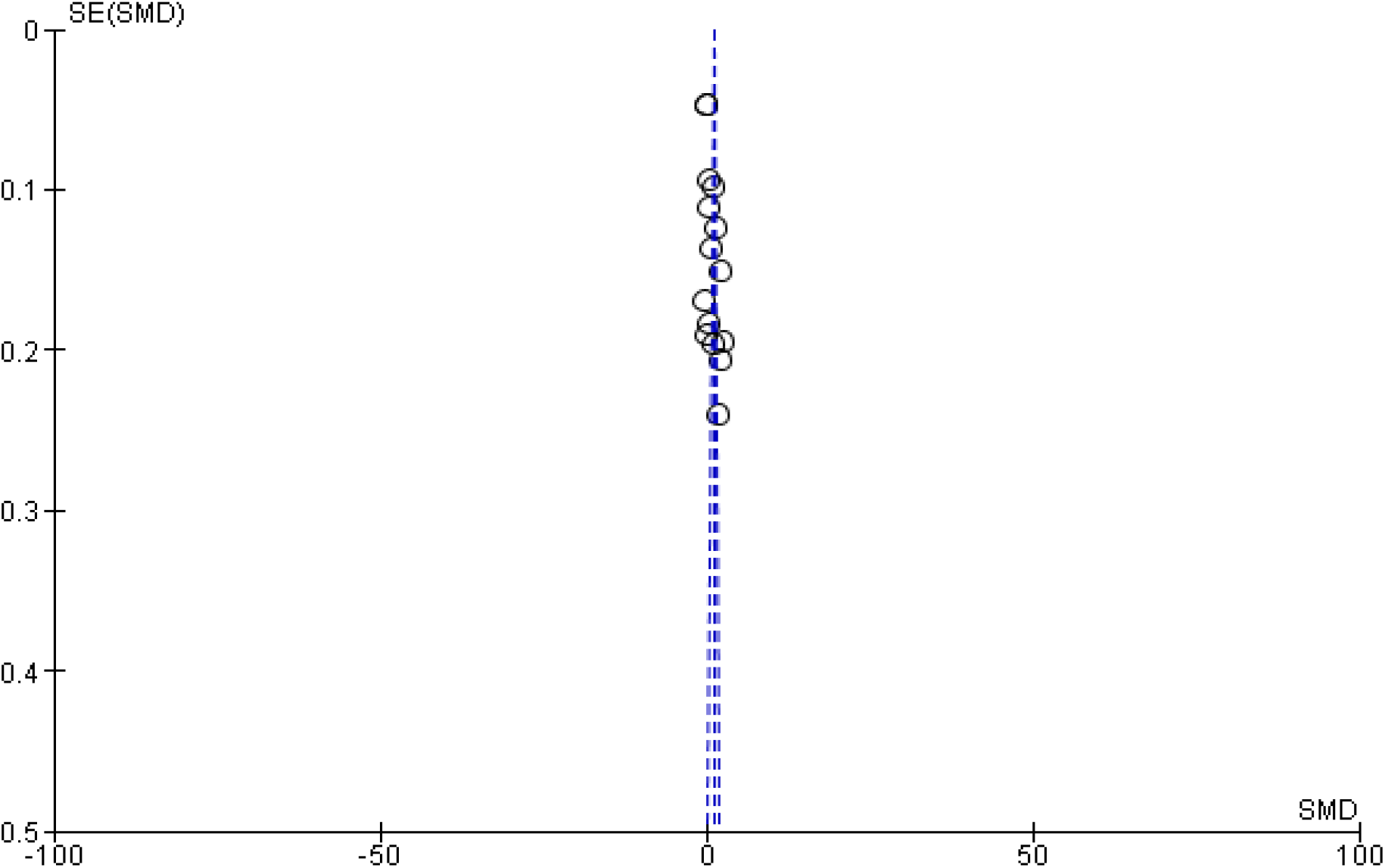
A funnel plot showing results of meta-analysis on the effects of maternal anemia interventions on hemoglobin levels. Data were reported as SMDs with 95% CIs.

To ascertain the highest possible effect of maternal anemia interventions, a further sensitivity analysis was conducted by categorically removing four studies[26], [27], [29], [31] (n= 2,116) whose individual interventions had shown no significant effect on hemoglobin levels: 0.18 [95% CI = −0.20, 0.55]; 0.08 [95% CI =−0.01, 0.17]; −0.29 [95% CI = −0.62, 0.04] and 0.28 [95% CI =−0.08, 0.64] respectively, there was a marginal change from the overall pooled effect [SMD (95% CI) = 0.93 [0.57, 1.30] to [SMD (95% CI) = 1.25 [0.77, 1.74] (n = 4,579) showing that, these studies’ interventions where compromised in one way or the other, by COVID-19 pandemic. Notably and surprisingly, these studies were conducted in developing continents with three of them [27], [29], [31] from Asia and one [26] from Africa (Fig 7).

**Figure 7.**
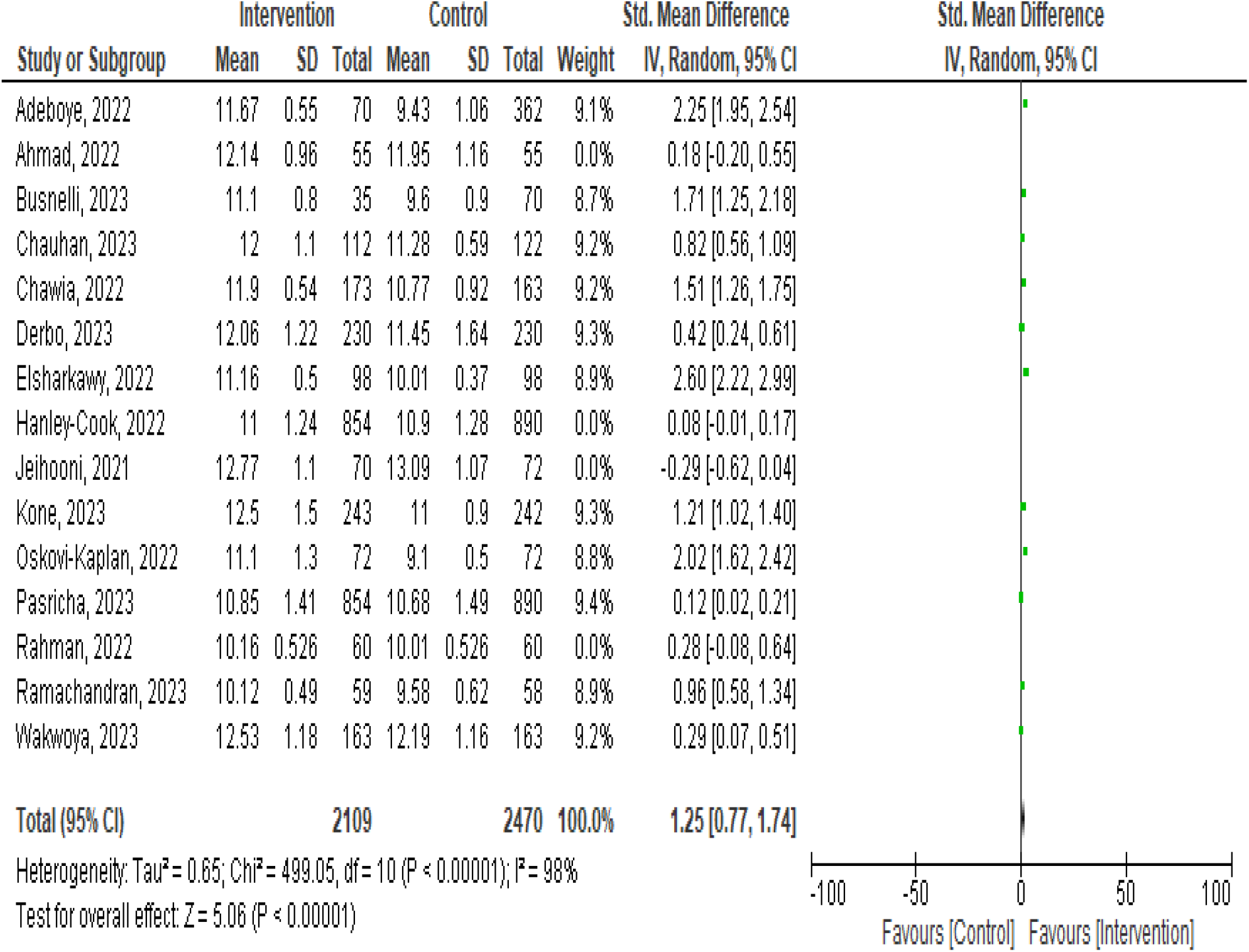
A forest plot showing results of sensitivity analysis after removing four studies (weight=0.0%)[26], [27], [29], [31] whose effects of maternal anemia interventions on hemoglobin levels had insignificant effect. Data were reported as MDs with 95% CIs.

## Discussion

The current systematic and meta-analysis study included 15 studies with 6,695 participants from different countries. The pooled cumulative effect of the maternal anemia interventions had a significant effect on the levels of serum hemoglobin level overall, with dietary iron supplementation intervention among pregnant women during COVID-19 showing no improvement on mean hemoglobin levels. Subgroup analysis propose that, parenterally given interventions greatly improved hemoglobin concentrations as opposed to orally administered interventions and interventions given via education package or information a significant utility on hemoglobin levels. However, with fixed effect model, a similar significant effect was found across all mode/routes of intervention administration. Sensitivity analysis for studies with low quality indicated and retained a substantial change in the effect of specific maternal anemia interventions on the mean hemoglobin levels noting that, studies conducted in developing countries majorly Asian, were the ones whose interventions did not bear any significant effect on hemoglobin levels as expected in control and treatment of maternal anemia. To the extent of our knowledge, this is the first meta-analysis which has evaluated and investigated an array of the overall effect of maternal anemia interventions on serum hemoglobin levels among pregnant cohort in the era of COVID-19.

According to existing evidence by some studies one being a meta-analysis like the current one [23], [38], [39], there was a positive association between specific form of maternal anemia intervention and serum hemoglobin concentration as it has been established in general from this current review. These studies collectively support the notion that targeted maternal interventions, such as nutrition education and counseling, can effectively improve hemoglobin levels and reduce the risk of anemia during pregnancy[40]. Notably, a meta-analysis study among pregnant women in 35 sub-Saharan African countries showed a 65.1% [95% CI = 64.9-65.3%] of non-adherence to iron supplementation during pandemics, a scenario that has been illuminated in the current study by demonstrating no improvement in serum hemoglobin levels probably, due to poor adherence on this dietary iron supplementation intervention. More explicitly to substantiate this current finding subject to dietary iron supplementation, a study conducted in Chile found that the use of iron supplements among pregnant women decreased during the pandemic, particularly among those with lower income and education levels[41]. Additionally, making reference to same study[41], the current finding indicated two studies [25], [26] that were based in Africa are the ones whose intervention category (dietary iron supplement) did not improve hemoglobin levels, may be due to lower income and education levels as a common issue in African nations. This study, conducted in 1, 507 pregnant women selected from public health care registries of the Southeast area of Santiago-Chile indicated that, non-supplement users before pregnancy had lower education and income.

Subject to parenterally given iron supplements in maternal anemia intervention showing more effect on hemoglobin levels compared to orally administered supplementation in the current study, this has been proven by another study that showed 87.8% of patients receiving intravenous iron had a significant rise in hemoglobin levels, compared to 65.85% of those receiving oral iron[42]. Interventions in form of education package and or information had a positive utility on hemoglobin levels in the current findings as it has been evidenced still in other findings that, the intervention group, which received trimester-based counseling sessions, regular SMS reminders, and educational leaflets on iron-rich diets, had a significant improvement in hemoglobin levels compared to the control group[23] and the intervention group received individualized nutrition education and a diet plan, while the control group received general education showing a significant increase in hemoglobin levels in the intervention group compared to the control group[43].

Previous studies have proposed that the COVID-19 pandemic significantly disrupted maternal health services in developing countries, including interventions aimed at reducing anemia and further showing that, the pandemic led to a decrease in family planning visits, antenatal and postnatal care visits, consultations for sick children, pediatric emergency visits, and child immunization levels compared to pre-pandemic levels. These disruptions likely affected the effectiveness of anemia interventions, as pregnant women had reduced access to essential healthcare services, including iron supplementation and nutritional counseling[44], [45]. The present meta-analysis has revealed similar results with studies conducted in developing countries, Asian [27], [29], [31] and African [26], having no effect on mean serum hemoglobin levels.

This meta-analysis had some limitations in that, from the funnel plot, even though the interventions’ effect of all the intervention aligned along the overall effect line, the studies were seemingly biased in publication. Dietary iron supplementation being the most common form of intervention, was only represented by two studies [25], [26]. This was though expected due to the compromising COVID-19 pandemic on access to such services[46]. Again, majority, almost half of studies[23], [27], [28], [29], [30], [31], [32] were based on education and information package as interventions rendered to pregnant women creating some form of imbalance however still, this is a demonstration that, it was the most feasible approach of mitigating maternal anemia in the era of during the COVID-19 pandemic, as providing education and information emerged as one of the most feasible ways to control maternal anemia with healthcare services disrupted and access to supplements limited, virtual counseling and educational interventions became crucial[47]. The possible reasons for the substantial heterogeneity in data among and between the included studies could be associated with the differences in the duration of supplementation, geographical location, sample size and the type of maternal anemia interventions. However, we performed the random-effects model, in order to determine the heterogeneity among these studies.

## Conclusion

In conclusion, the current systematic review and meta-analysis demonstrated that generally, maternal anemia interventions improved the serum hemoglobin level mostly, the parenterally (injected) given. Dietary iron supplements as maternal anemia intervention didn’t improve while carboxyl-maltose had the most superior effect on improving serum hemoglobin level. Interventions from developing countries were highly compromised by COVID-19 pandemic with no impact on hemoglobin levels. The current study suggests that parenterally given maternal anemia interventions as opposed to oral given ones are more feasible in improving serum hemoglobin levels in such a compromising event of a pandemic. Further tailored approaches are crucial in developing nations with possible fragile healthcare systems in regards to maternal anemia interventions and that, most convenient way of intervention across board could be pertinent information and educating the pregnant women is such situations. Therefore, further high-quality, well designed and long term RCTs in this field are extensively required.

## Data Availability

All data produced in the present study are available upon reasonable request to the authors

## Notes

### Competing Interest Statement

The authors have declared no competing interest.

### Funding Statement

Partially by KMTC, Kenya

